## Supplementary Information for "Strength and durability of indirect protection against SARS-CoV-2 infection through vaccine and infection-acquired immunity"

### Contents

Supplementary Notes……………………………………………………………………...2
References…………………………………………………………………………………5
Supplementary Tables……………………………………………………………………..6
Supplementary Figures…………………………………………………………….…..21

### Supplementary Notes

*SARS-CoV-2 surveillance program in California prison system*
During the COVID-19 pandemic, the California state prison system implemented a SARS-CoV-2 public health surveillance program with frequent systematic viral testing to improve case detection and reduce transmission through early isolation. This testing included routine asymptomatic SARS-CoV-2 viral testing (e.g., weekly testing), reactive asymptomatic testing during periods of high transmission (e.g., once, weekly, after exposure), symptomatic testing, and testing for routine clinical care[1,3].

Testing was optional, but testing rates remained broadly consistent over time (Figure 1). Test acceptance was high during our study period; approximately 70% of residents participated in some testing between December 15, 2021, and December 15, 2022.

The predominant test used was PCR (78% of tests were PCR during the study period), which often had a 1-5 day delay from sample collection to test result. This delay in PCR result was common in all settings during this period, including in the general US population. Some testing was performed with rapid antigen tests which provided same day results. Turnaround time for test results during our study period was the same day for 30% of tests, one day for 37% of tests, and 2 or more days for 33% of tests.

*Isolation and quarantine policies*
Isolation and quarantine policy was set by the California Correctional Health Care Services. During the study period, residents that tested positive for SARS-CoV-2 were moved into isolation for 10-14 days upon receiving the positive test result, either in a single room or group isolation with others with active SARS-CoV-2 infection. Residents were placed in quarantine if they had a confirmed or suspected SARS-CoV-2 exposure[2]. For example, 25% of residents with a new SARS-CoV-2 infection in the study period were placed in quarantine (resided alone) up to 3 days before their first positive test (based on a prior exposure). Lockdowns (where movement was restricted) occurred at a building level if three or more cases were detected in a building within 14 days.

*COVID-19 vaccination*
COVID-19 vaccines became available to residents in the California state prison system in December 2020. Most residents received mRNA vaccines, including BNT162b2 (65%) and mRNA-1273 (18%), as their primary COVID-19 vaccine series. Only 17% of residents received the Ad26.COV2.S vaccine. In our final study population, 6% of cases and controls and 5% of their roommates received the Ad26.COV2.S vaccine. For this analysis, we assumed residents were fully vaccinated after receiving 2 doses of a mRNA vaccine or one dose of the Ad26.COV2.S vaccine.

*Mechanism of indirect protection*
The expected mechanism for indirect protection is that vaccine-derived and/or infection-acquired immunity in roommates reduces their risk of SARS-CoV-2 infection and/or their infectiousness with a SARS-CoV-2 infection, thereby reducing the risk of SARS-CoV-2 transmission to the case or control. To further evaluate this mechanism empirically within our study population, we

analyzed testing data of roommates in the 14 and 4 days prior to test collection in cases and controls (Supplementary Table 1). Test acceptance in the 14 days prior to study inclusion among roommates of cases and controls was similar, suggesting similar testing patterns. Roommates of cases were more likely to have new SARS-CoV-2 infection within 4 days of test collection than roommates of controls, suggesting within room transmission and plausibility for indirect protection at a room level.

*Additional notes on matching*

Matching of cases and controls included both exact and distance-based matching. We matched cases and controls in a 1:2 ratio exactly by time (test collection within 2 days), COVID-19 vaccine status (by dose), prior SARS-CoV-2 infection, and building and security level. Cases and controls were eligible for matching if they were not roommates. Cases and controls were then matched to minimize the Euclidean distance based on age and risk score for severe COVID-19 for both the case or control and their roommate and the time (days) since last COVID-19 vaccine and/or SARS-CoV-2 infection of the case or control. All distance-based variables were standardized, and time since last COVID-19 vaccine and time since most recent SARS-CoV-2 infection were weighted 2x in the distance score based on their importance for matching.

Cases that were matched with multiple negative tests (controls) from the same resident that met study and matching criteria were rematched so residents would not be repeated within a matched group. Not all cases were matched to two controls; approximately 30% of cases had only one matched control. Match quality is shown in Supplementary Figure 1.

*Additional notes on statistical analysis and data*

The analysis was a test-negative case-control design with measurement taken from the study population. Residents were eligible for inclusion throughout the study period based on their testing data; however, residents were not included more than once within a matched group. Of the 9,625 residents included in the study, most were included only once as a case or control (N=7,555), and 2,070 residents were included more than once as a case and/or control in distinct matched groups. Each observation of a resident was treated independently in the main analysis since a minority of individuals were repeated and the within person correlation may be less significant in this analysis. However, to evaluate this assumption, we assessed the sensitivity of our results to repeated measures in multiple sensitivity analyses (Supplementary Table 15), including removal of repeated residents. Case and controls missing any covariate data (security level, age, and risk score for severe COVID-19) (<0.1%) were removed prior to matching. Statistical tests are two-sided.

*Additional notes on analysis of bivalent vaccines*

Ancestral monovalent vaccines were administered in the study population through August 2022. In September 2022, CCHCS began to administer bivalent vaccines, which targeted both ancestral SARS-CoV-2 and Omicron subvariants BA.4 and BA.5. We conducted an analysis that examined the indirect protection from bivalent COVID-19 vaccines (administered after September 1, 2022). We defined vaccine exposure as a categorical variable (unvaccinated, vaccinated with ancestral monovalent dose within 3 months, vaccinated with ancestral monovalent dose more than three months ago, and vaccinated with bivalent dose) to estimate indirect protection from both ancestral monovalent vaccines and bivalent vaccines against Omicron SARS-CoV-2 infection. Therefore, the bivalent vaccine analysis was limited to September – December 2022; in contrast, the ancestral monovalent vaccine analysis was during the period before bivalent vaccine introduction (December 2021 – August 2022). We recognize two key limitations in this analysis: i) vaccines were administered at different time points during the study period limiting comparability, although we adjust for many potential confounders; and ii) our estimates of bivalent vaccine indirect protection are underpowered and have a limited follow up period.

*Pre-analysis plan*
The pre-analysis plan is available here: https://github.com/sophttan/covid-indirects. The final primary analysis had minor changes from the initial plan. We added an additional requirement that controls did not have a positive test in the 14 days after collection of their negative test to ensure these controls were truly negative. We tested multiple definitions for timing of co-residence of cases and controls with their roommates to account for the latent period between exposure and infection and movement of residents (quarantine after exposure). To further improve match quality, we matched cases and controls by time since their most recent vaccine and/or infection (in addition to number of vaccine doses and binary prior infection status); we also report results without these additional matching criteria.

*IRB approval*
This study was approved by the Institutional Review Board (IRB) at Stanford University (#70927) and the University of California, San Francisco (#21-34030). The study was retrospective using secondary data without direct identifiers collected for operational purposes. The IRB included a waiver of consent based on the following criteria: deemed no more than minimal risk to subjects; could not practicably be done without the waiver; could not practicably be done without identifiable information; will not adversely affect rights and welfare of subjects; and will provide subjects with additional pertinent information after participation[3].

### Supplementary Tables

#### Supplementary Table 1. Testing and SARS-CoV-2 infections in cases, controls, and their roommates prior to test collection in cases and controls.

| | | Case/Control | | | | Roommates | | | |
|---|---|---|---|---|---|---|---|---|---|
| | | Case | | Control | | Case | | Control | |
| Before test collection | Vaccine or prior infection | N | n (%) | N | n (%) | N | n (%) | N | n (%) |
| **Any SARS-CoV-2 test within 90 days** | None | 331 | 293 (89%) | 484 | 415 (86%) | 388 | 327 (84%) | 487 | 417 (86%) |
| | Any | 4,309 | 3,747 (87%) | 7,340 | 6,449 (88%) | 4,252 | 3,713 (87%) | 7,337 | 6,447 (88%) |
| **Any SARS-CoV-2 test within 14 days** | None | 331 | 191 (58%) | 484 | 284 (59%) | 388 | 218 (56%) | 487 | 275 (57%) |
| | Any | 4,309 | 2,628 (61%) | 7,340 | 4,615 (63%) | 4,252 | 2,601 (61%) | 7,337 | 4,624 (63%) |
| **Any SARS-CoV-2 test within 4 days** | None | 331 | 53 (16%) | 484 | 66 (14%) | 388 | 69 (18%) | 487 | 79 (16%) |
| | Any | 4,309 | 779 (18%) | 7,340 | 1,283 (20%) | 4,252 | 763 (18%) | 7,337 | 1,470 (20%) |
| **SARS-CoV-2 infection within 4 days** | None | - | - | - | - | 69 | 47 (68%) | 79 | 24 (30%) |
| | Any | - | - | - | - | 763 | 315 (41%) | 1,470 | 211 (14%) |

We compared testing practices and SARS-CoV-2 infections in the cases/controls and their roommates. We examined testing of cases, controls, and roommates at 4, 14, and 90 days prior to test collection in the cases and controls to ensure similar testing practices. We evaluated the mechanism of indirect protection, which is reduced risk of SARS-CoV-2 infection or infectiousness in the roommate due to their vaccination and/or infection-acquired immunity (thus yielding indirect protection), by comparing new infections in roommates of cases versus controls in the 4 days before test collection (in the subset with available testing data).

**Supplementary Table 2. Primary model results of indirect protection from vaccine-derived and infection-acquired immunity.**

| | Odds ratio (OR) (95% CI) |
|---|---|
| **Vaccine-derived immunity (any)** | 0.778 (0.694, 0.872) |
| **Infection-acquired immunity (any)** | 0.841 (0.773, 0.915) |
| **Age** | |
| Case or control | 0.995 (0.99, 1.00) |
| Roommate | 1.00 (1, 1.01) |
| **Severe COVID-19 risk score** | |
| Case or control | 1.08 (1.04, 1.11) |
| Roommate | 1.08 (1.04, 1.11) |

We fit a conditional logistic regression model to estimate the indirect protection against Omicron SARS-CoV-2 infection conferred to individuals with a roommate with any vaccine-derived and/or any infection-acquired immunity (prior SARS-CoV-2 infection). Indirect protection was estimated as 1-OR from the adjusted model. We adjusted for age and risk of severe COVID-19 of both the case/control and their roommate.

**Supplementary Table 3. Vaccine dose-dependent indirect protection from vaccine-derived immunity.**

| | Odds ratio (OR) (95% CI) | |
|---|---|---|
| | Dose (numeric) | Dose (categorical) |
| **Vaccine-derived immunity (by dose)** | | |
| Partial vaccination | 0.925 (0.891, 0.96) | 0.584 (0.408, 0.836) |
| Primary series alone | 0.856 (0.824, 0.888) | 0.817 (0.715, 0.933) |
| One booster | 0.791 (0.763, 0.821) | 0.778 (0.690, 0.878) |
| Two or more boosters | 0.732 (0.705, 0.76) | 0.698 (0.571, 0.852) |
| **Infection-acquired immunity (any)** | 0.839 (0.771, 0.913) | 0.840 (0.772, 0.914) |
| **Age** | | |
| Case or control | 0.995 (0.99, 1.00) | 0.995 (0.99, 1.00) |
| Roommate | 1.01 (1.00, 1.01) | 1.01 (1.00, 1.01) |
| **Severe COVID-19 risk score** | | |
| Case or control | 1.08 (1.04, 1.11) | 1.08 (1.04, 1.11) |
| Roommate | 1.08 (1.05, 1.11) | 1.08 (1.05, 1.12) |

We fit a conditional logistic regression model to estimate the vaccine dose-dependent indirect protection against Omicron SARS-CoV-2 infection conferred to individuals with a roommate with vaccine-derived and/or infection-acquired immunity. In our main analysis, vaccination was a numeric variable that represented the following vaccine statuses: unvaccinated, partial vaccination with mRNA vaccine, primary series alone, one booster dose, and two or more booster doses. We also report a sensitivity analysis that defined vaccination as a categorical variable, and results are similar (estimates of indirect protection from partially vaccinated roommates had a limited sample size). Indirect protection was estimated as 1-OR from the adjusted model. We adjusted for age and risk of severe COVID-19 of both the case/control and their roommate and for the prior infection status of the roommate.

**Supplementary Table 4. Indirect protection from vaccine-derived, infection-acquired, and hybrid immunity.**

| | Odds ratio (OR) (95% CI) |
|---|---|
| **Immunity** | |
| Only vaccine-derived immunity | 0.752 (0.646, 0.876) |
| Only infection-acquired immunity | 0.787 (0.634, 0.977) |
| Both vaccine- and infection-acquired immunity (hybrid) | 0.638 (0.544, 0.749) |
| **Age** | |
| Case or control | 0.995 (0.99, 1.00) |
| Roommate | 1.00 (1, 1.01) |
| **Severe COVID-19 risk score** | |
| Case or control | 1.08 (1.04, 1.12) |
| Roommate | 1.09 (1.05, 1.12) |

To directly evaluate the indirect protection from co-residing with an individual with both vaccine-derived and infection-acquired immunity (hybrid immunity), we grouped vaccine and infection exposures (no vaccine- or infection-acquired immunity, only vaccine-derived immunity, only infection-acquired immunity, and both vaccine- and infection-acquired immunity) and used conditional logistic regression models to quantify the indirect protection against Omicron SARS-CoV-2 infection. Indirect protection was estimated as 1-OR from the adjusted model. We adjusted for age and risk of severe COVID-19 of both the case/control and their roommate.

**Supplementary Table 5. Temporal dynamics of indirect protection since most recent vaccination, most recent SARS-CoV-2 infection, and most recent vaccine or infection.**

| | Odds ratio (OR) (95% CI) based on time since most recent vaccine and/or infection | | |
|---|---|---|---|
| | **Vaccine** | **Infection** | **Vaccine or infection** |
| **Vaccine-derived immunity (time)** | | | |
| <3 months | 0.703 (0.619, 0.798) | 0.628 (0.451, 0.875) | 0.639 (0.545, 0.749) |
| 3 to <6 months | 0.871 (0.749, 1.01) | 0.616 (0.476, 0.796) | 0.757 (0.635, 0.903) |
| 6 to <12 months | 0.830 (0.727, 0.949) | 0.746 (0.619, 0.898) | 0.774 (0.658, 0.912) |
| 12+ months | 0.820 (0.671, 1.00) | 0.884 (0.808, 0.967) | 0.822 (0.675, 1.00) |
| **Vaccine-derived immunity (any)** | - | 0.768 (0.685, 0.861) | - |
| **Infection-acquired immunity (any)** | 0.834 (0.767, 0.908) | - | - |
| **Age** | | | |
| Case or control | 0.995 (0.99, 1.00) | 0.995 (0.990, 1.00) | 0.995 (0.99, 1.00) |
| Roommate | 1.01 (1.00, 1.01) | 1.00 (0.999, 1.01) | 1.01 (1.00, 1.01) |
| **Severe COVID-19 risk score** | | | |
| Case or control | 1.08 (1.04, 1.11) | 1.08 (1.04, 1.11) | 1.08 (1.04, 1.11) |
| Roommate | 1.08 (1.04, 1.11) | 1.08 (1.04, 1.11) | 1.08 (1.04, 1.11) |

We fit three separate conditional logistic regression models to estimate the indirect protection against Omicron SARS-CoV-2 infection in individuals based on the time since their roommate's most recent vaccination, SARS-CoV-2 infection, or most recent event (vaccine dose or infection). Time since event begins 14 days after the event to reflect lag in the onset of protection. Indirect protection was estimated as 1-OR from the adjusted model. We adjusted for age and risk of severe COVID-19 of both the case/control and their roommate in each model.

**Supplementary Table 6. Indirect protection in the first three months of vaccine-derived immunity.**

| Time since vaccine | Odds ratio (OR) (95% CI) |
|---|---|
| <1 months | 0.650 (0.547, 0.773) |
| 1 to <2 months | 0.693 (0.597, 0.804) |
| 2 to <3 months | 0.773 (0.655, 0.912) |

Since vaccine-derived protection is strongest within the first three months of vaccine-derived immunity, we fit a conditional logistic regression model to estimate the indirect protection against Omicron SARS-CoV-2 infection conferred to individuals from their roommates within the first three months of vaccination. Time since vaccine begins 14 days after vaccine receipt to reflect lag in the onset of protection. Indirect protection was estimated as 1-OR from the adjusted model. We adjusted for age and risk of severe COVID-19 of both the case/control and their roommate and for the prior infection status of the roommate.

**Supplementary Table 7. Indirect protection estimated by time since most recent vaccine and most recent infection.**

| | Odds ratio (OR) (95% CI) |
|---|---|
| **Vaccine-derived immunity (time)** | |
| <3 months | 0.694 (0.611, 0.788) |
| 3 to <6 months | 0.856 (0.736, 0.995) |
| 6 to <12 months | 0.821 (0.719, 0.939) |
| 12+ months | 0.816 (0.667, 0.998) |
| **Infection-acquired immunity (time)** | |
| <3 months | 0.621 (0.446, 0.866) |
| 3 to <6 months | 0.612 (0.473, 0.792) |
| 6 to <12 months | 0.743 (0.617, 0.895) |
| 12+ months | 0.877 (0.802, 0.96) |

We fit a single conditional logistic regression model to estimate the indirect protection against Omicron SARS-CoV-2 infection in individuals based on time since their roommate's most recent vaccination and/or most recent SARS-CoV-2 infection. Results are similar to the main models where we fit separate models and adjusted for binary vaccination or infection. Time since event begins 14 days after the event to reflect lag in the onset of protection. Indirect protection was estimated as 1-OR from the adjusted model. We adjusted for age and risk of severe COVID-19 of both the case/control and their roommate.

### Supplementary Table 8. Indirect protection from infection-acquired immunity from pre-Omicron infection and Omicron infection.

| | pre-Omicron infection | | Omicron infection | |
|---|---|---|---|---|
| **Infection-acquired immunity** | **N** | **Odds ratio (OR) (95% CI)** | **N** | **Odds ratio (OR) (95% CI)** |
| **Time** | | | | |
| <3 months | 4 | - | 192 | 0.631 (0.451, 0.883) |
| 3 to <6 months | 51 | 0.624 (0.323, 1.20) | 310 | 0.614 (0.464, 0.811) |
| 6 to <12 months | 234 | 0.783 (0.5574, 1.07) | 482 | 0.728 (0.580, 0.913) |
| 12+ months | 4,933 | 0.884 (0.808, 0.967) | - | - |

Since indirect protection from infection-acquired immunity may be different over time against different variants, we estimated indirect protection when we categorized prior infection by variant (pre-Omicron v. Omicron) and by time since infection. Indirect protection was estimated as 1-OR from the adjusted model. We adjusted for age and risk of severe COVID-19 of both the case/control and their roommate and vaccine-derived immunity (any) in the roommate. We show the sample size within each category as well as the estimated odds ratio of Omicron SARS-CoV-2 infection. This analysis is underpowered (especially for pre-Omicron infection <3 and 3-6 months), though point estimates of indirect protection within 6-12 months of pre-Omicron and Omicron infection are similar.

**Supplementary Table 9. Interaction between indirect protection from vaccine-derived and infection-acquired immunity.**

| Odds ratio (OR) (95% CI) | | Infection-acquired immunity | | | | |
|---|---|---|---|---|---|---|
| | | **Any** | **<3 months** | **3 to <6 months** | **6 to <12 months** | **12+ months** |
| **Vaccine-derived immunity** | **Any** | 1.08 (0.86, 1.35) | 0.92 (0.391, 2.16) | 0.922 (0.480, 1.77) | 1.05 (0.691, 1.6) | 1.08 (0.835, 1.39) |
| | **<3 months** | 1.07 (0.836, 1.36) | 0.867 (0.325, 2.31) | 0.720 (0.325, 1.59) | 1.01 (0.608, 1.69) | 1.06 (0.810, 1.39) |
| | **3 to <6 months** | 1.09 (0.820, 1.46) | 1.44 (0.481, 4.30) | 1.16 (0.537, 2.53) | 1.27 (0.688, 2.34) | 1.05 (0.763, 1.43) |
| | **6 to <12 months** | 1.04 (0.8, 1.35) | 0.669 (0.252, 1.77) | 0.865 (0.407, 1.84) | 0.958 (0.591, 1.55) | 1.09 (0.813, 1.45) |
| | **12+ months** | 1.13 (0.768, 1.65) | 1.86 (0.360, 9.56) | 0.922 (0.368, 2.31) | 0.968 (0.510, 1.84) | 1.27 (0.819, 1.97) |

We tested for multiplicative interactions between vaccine-derived and infection-acquired indirect protection against Omicron SARS-CoV-2 infection. We fit four conditional logistic regression models to assess the interactions between: 1) any vaccine-derived immunity and any infection-acquired immunity (blue); 2) vaccine-derived immunity by time and any infection-acquired immunity (orange); 3) any vaccine-derived immunity and infection-acquired immunity by time (green); and 4) vaccine-derived immunity by time and infection-acquired immunity by time (gray). Time since vaccine begins 14 days after vaccine receipt to reflect lag in the onset of protection. Indirect protection was estimated as 1-OR from the adjusted model. We adjusted for age and risk of severe COVID-19 of both the case/control and their roommate and for the prior infection status of the roommate.

**Supplementary Table 10. Indirect protection from vaccine-derived and infection-acquired immunity stratified by immunity of matched cases and controls.**

| | Odds ratio (OR) (95% CI) | | | | |
|---|---|---|---|---|---|
| | | Cases and controls with different immunity status | | | |
| | All cases and controls (main) | No immunity (N=815) | Only vaccine-derived immunity (N=6,225) | Only infection-acquired immunity (N=543) | Both vaccine- and infection-acquired immunity (hybrid) (N=4,881) |
| **Vaccine-derived immunity** | | | | | |
| Any | 0.778 (0.694, 0.872) | 0.758 (0.551, 1.04) | 0.759 (0.643, 0.896) | 1.04 (0.666, 1.61) | 0.757 (0.619, 0.925) |
| By dose | | | | | |
| Partially vaccinated | 0.925 (0.891, 0.96) | 0.899 (0.798, 1.01) | 0.918 (0.87, 0.968) | 1.02 (0.872, 1.19) | 0.926 (0.870, 0.985) |
| Primary series only | 0.856 (0.824, 0.888) | 0.808 (0.718, 0.910) | 0.842 (0.798, 0.889) | 1.04 (0.889, 1.21) | 0.857 (0.806, 0.912) |
| One booster | 0.791 (0.763, 0.821) | 0.727 (0.645, 0.818) | 0.773 (0.732, 0.816) | 1.06 (0.906, 1.24) | 0.793 (0.746, 0.844) |
| Two or more boosters | 0.732 (0.705, 0.76) | 0.653 (0.580, 0.736) | 0.71 (0.672, 0.749) | 1.08 (0.923, 1.26) | 0.734, 0.691, 0.781) |
| By time | | | | | |
| <3 months | 0.703 (0.619, 0.798) | 0.704 (0.480, 1.03) | 0.686 (0.571, 0.824) | 0.964 (0.576, 1.61) | 0.672 (0.538, 0.838) |
| 3-6 months | 0.871 (0.749, 1.01) | - | 0.817 (0.658, 1.01) | - | 0.973 (0.757, 1.25) |
| 6-12 months | 0.830 (0.727, 0.949) | 0.925 (0.608, 1.41) | 0.831 (0.684, 1.01) | 1.06 (0.617, 1.81) | 0.767 (0.611, 0.962) |
| 12+ months | 0.820 (0.671, 1.00) | - | 0.828 (0.618, 1.11) | - | 0.712 (0.514, 0.987) |
| **Infection-acquired immunity** | | | | | |
| Any | 0.841 (0.773, 0.915) | 0.837 (0.583, 1.20) | 0.806 (0.715, 0.909) | 0.821 (0.558, 1.21) | 0.902 (0.788, 1.03) |
| By time | | | | | |
| <3 months | 0.628 (0.451, 0.875) | - | - | - | 0.799 (0.512, 1.25) |
| 3-6 months | 0.616 (0.476, 0.796) | - | 0.702 (0.479, 1.03) | - | 0.662 (0.450, 0.973) |
| 6-12 months | 0.746 (0.619, 0.898) | - | 0.719 (0.540, 0.958) | - | 0.724 (0.547, 0.959) |
| 12+ months | 0.884 (0.808, 0.967) | 0.818 (0.547, 1.22) | 0.841 (0.739, 0.956) | 0.913 (0.610, 1.37) | 0.952 (0.826, 1.1) |

We conducted a sub-analysis to evaluate differences in indirect protection based on immunity status in cases and controls. We used separate conditional logistic regression models to quantify indirect protection against Omicron SARS-CoV-2 infection among cases and controls in four groups: no immunity, vaccine-derived immunity only, infection-acquired immunity only, and hybrid immunity. Indirect protection was estimated as 1-OR from the adjusted model. We adjusted for age and risk of severe COVID-19 of both the case/control and their roommate. The sample size was highly variable between sub-groups, and some analyses were underpowered due to sample size. The indirect protection results are similar across sub-groups, with the exception of infection-acquired immunity, although the small sample size here limits interpretation. Results for sub-analyses where sample size was less than 100 cases and controls are not reported.

**Supplementary Table 11. Indirect protection from bivalent vaccine.**

| | Ancestral monovalent vaccine | | | Bivalent vaccine | | |
|---|---|---|---|---|---|---|
| **Time since vaccine** | Average time since vaccine (days) | N | Odds ratio (OR) (95% CI) | Average time since vaccine (days) | N | Odds ratio (OR) (95% CI) |
| <3 months | 48 | 5,155 | 0.697 (0.613, 0.792) | 22 | 122 | 0.597 (0.367, 0.973) |
| 3+ months | 243 | 5,638 | 0.831 (0.735, 0.939) | - | - | - |

We estimated the indirect protection from ancestral monovalent vaccines (administered December 2021 – August 2022) and the new bivalent vaccine formulation (administered starting September 2022). Vaccination was categorized: unvaccinated, vaccinated with ancestral monovalent dose within 3 months, vaccinated with ancestral monovalent dose more than three months ago, and vaccinated with bivalent dose (received on or after September 1, 2022). No bivalent vaccines were received more than 3 months prior to the end of the study. We fit a conditional logistic regression model to estimate the indirect protection against Omicron SARS-CoV-2 infection from roommates. Time since event begins 14 days after vaccine receipt to reflect lag in the onset of protection. Indirect protection was estimated as 1-OR from the adjusted model. We adjusted for age and risk of severe COVID-19 of both the case/control and their roommate and for the prior infection status of the roommate. On average, bivalent vaccines were received more recently than the ancestral vaccine within the <3 month group.

**Supplementary Table 12. Negative control exposure analysis to estimate indirect protection against SARS-CoV-2 infection from influenza vaccination in roommates.**

| | Percentage of roommates with influenza vaccination | | |
|---|---|---|---|
| | Case roommates | Control roommates | **Odds ratio (OR) (95% CI)** |
| Any influenza vaccine (lifetime) | 78% | 77% | 1.05 (0.955, 1.15) |
| Influenza vaccine within 2 years | 64% | 64% | 1.01 (0.929, 1.09) |
| Influenza vaccine within 1 year | 52% | 52% | 0.994 (0.918, 1.08) |
| Time since influenza vaccine (months) | - | - | 1.00 (0.998, 1.00) |

We estimated the indirect protection against Omicron SARS-CoV-2 infection conferred by influenza vaccination in the roommate to cases and controls as a negative control analysis. We fit separate conditional logistic regression models with the same study population of matched cases and controls where the primary exposures were different influenza vaccine statuses of roommates. The null results argue against substantial residual confounding.

**Supplementary Table 13. Sensitivity analyses of indirect protection from vaccine and infection-acquired immunity to different definition of co-residence in housing for cases, controls, and their roommates.**

| | Odds ratio (OR) (95% CI)<br>Roommate defined based on different time periods prior to test collection | | | |
|---|---|---|---|---|
| | **Days 3-6 (main)** | **Day 3** | **Days 0-6** | **Days 6-9** |
| **Vaccine-derived immunity** | | | | |
| Any | 0.778 (0.694, 0.872) | 0.775 (0.694, 0.865) | 0.776 (0.689, 0.874) | 0.785 (0.703, 0.876) |
| By dose | | | | |
| Partially vaccinated | 0.925 (0.891, 0.96) | 0.924 (0.892, 0.958) | 0.929 (0.894, 0.965) | 0.930 (0.898, 0.964) |
| Primary series only | 0.856 (0.824, 0.888) | 0.854 (0.824, 0.885) | 0.863 (0.830, 0.897) | 0.866 (0.835, 0.897) |
| One booster | 0.791 (0.763, 0.821) | 0.78 (0.762, 0.818) | 0.801 (0.771, 0.833) | 0.805 (0.777, 0.835) |
| Two or more boosters | 0.732 (0.705, 0.76) | 0.73 (0.704, 0.757) | 0.744 (0.716, 0.774) | 0.749 (0.723, 0.777) |
| By time | | | | |
| <3 months | 0.703 (0.619, 0.798) | 0.712 (0.630, 0.805) | 0.707 (0.619, 0.806) | 0.710 (0.628, 0.803) |
| 3-6 months | 0.871 (0.749, 1.01) | 0.848 (0.734, 0.979) | 0.875 (0.748, 1.02) | 0.867 (0.75, 1.00) |
| 6-12 months | 0.830 (0.727, 0.949) | 0.816 (0.718, 0.927) | 0.82 (0.714, 0.942) | 0.849 (0.746, 0.965) |
| 12+ months | 0.820 (0.671, 1.00) | 0.817 (0.674, 0.990) | 0.821 (0.667, 1.01) | 0.801 (0.659, 0.975) |
| **Infection-acquired immunity** | | | | |
| Any | 0.841 (0.773, 0.915) | 0.841 (0.775, 0.912) | 0.882 (0.808, 0.963) | 0.847 (0.779, 0.92) |
| By time | | | | |
| <3 months | 0.628 (0.451, 0.875) | 0.665 (0.499, 0.886) | 0.688 (0.489, 0.969) | 0.738 (0.514, 1.06) |
| 3-6 months | 0.616 (0.476, 0.796) | 0.656 (0.513, 0.839) | 0.659 (0.504, 0.861) | 0.631 (0.491, 0.812) |
| 6-12 months | 0.746 (0.619, 0.898) | 0.805 (0.673, 0.963) | 0.767 (0.634, 0.928) | 0.724 (0.603, 0.869) |
| 12+ months | 0.884 (0.808, 0.967) | 0.872 (0.8, 0.951) | 0.928 (0.846, 1.02) | 0.888 (0.814, 0.969) |

In our primary analysis, we required cases and controls to co-reside with a single resident in the 3-6 days leading up to test collection and study inclusion. This accounted for the latent period between exposure and detectable infection, which is relevant to this study on indirect protection provided by the roommate's vaccine-derived or infection-acquired immunity; additionally, this accounted for movement of residents for quarantine due to exposure. We tested alternative definitions of this housing requirement, including co-residing together on the 3rd day prior to test collection, the week leading up to test collection, and 6-9 days prior to test collection. All other study design remained the same, including matching and statistical analysis.

**Supplementary Table 14. Sensitivity analyses of indirect protection from vaccine and infection-acquired immunity with different matching and variable specifications.**

| | Odds ratio (OR) (95% CI) | | | | |
|---|---|---|---|---|---|
| | **Main** | **1:1 matching** | **Removing matching by time since vaccine and/or infection** | **Adding statistical adjustment for time since vaccine and/or infection of case/control** | **Removing statistical adjustment for age and risk** |
| **Vaccine-derived immunity** | | | | | |
| Any | 0.778 (0.694, 0.872) | 0.798 (0.703, 0.906) | 0.784 (0.699, 0.879) | 0.773 (0.689, 0.866) | 0.791 (0.706, 0.886) |
| By dose | | | | | |
| Partially vaccinated | 0.925 (0.891, 0.96) | 0.93 (0.892, 0.969) | 0.927 (0.893, 0.963) | 0.923 (0.9, 0.958) | 0.943 (0.909, 0.978) |
| Primary series only | 0.856 (0.824, 0.888) | 0.865 (0.83, 0.901) | 0.860 (0.829, 0.893) | 0.853 (0.821, 0.885) | 0.889 (0.857, 0.922) |
| One booster | 0.791 (0.763, 0.821) | 0.804 (0.772, 0.839) | 0.798 (0.769, 0.828) | 0.787 (0.759, 0.817) | 0.838 (0.808, 0.869) |
| Two or more boosters | 0.732 (0.705, 0.76) | 0.748 (0.718, 0.779) | 0.74 (0.713, 0.768) | 0.727 (0.700, 0.755) | 0.790 (0.762, 0.82) |
| By time | | | | | |
| <3 months | 0.703 (0.619, 0.798) | 0.730 (0.634, 0.841) | 0.707 (0.623, 0.804) | 0.700 (0.617, 0.796) | 0.732 (0.646, 0.830) |
| 3-6 months | 0.871 (0.749, 1.01) | 0.892 (0.755, 1.05) | 0.87 (0.748, 1.01) | 0.867 (0.746, 1.01) | 0.907 (0.781, 1.05) |
| 6-12 months | 0.830 (0.727, 0.949) | 0.834 (0.719, 0.968) | 0.838 (0.733, 0.958) | 0.821 (0.719, 0.939) | 0.817 (0.716, 0.933) |
| 12+ months | 0.820 (0.671, 1.00) | 0.852 (0.681, 1.07) | 0.849 (0.694, 1.04) | 0.815 (0.665, 0.999) | 0.796 (0.652, 0.973) |
| **Infection-acquired immunity** | | | | | |
| Any | 0.841 (0.773, 0.915) | 0.826 (0.752, 0.906) | 0.833 (0.766, 0.906) | 0.843 (0.775, 0.918) | 0.838 (0.77, 0.886) |
| By time | | | | | |
| <3 months | 0.628 (0.451, 0.875) | 0.556 (0.388, 0.798) | 0.633 (0.454, 0.883) | 0.632 (0.453, 0.882) | 0.632 (0.454, 0.879) |
| 3-6 months | 0.616 (0.476, 0.796) | 0.639 (0.480, 0.852) | 0.589 (0.457, 0.758) | 0.636 (0.490, 0.825) | 0.604 (0.467, 0.781) |
| 6-12 months | 0.746 (0.619, 0.898) | 0.750 (0.609, 0.924) | 0.717 (0.596, 0.861) | 0.761 (0.631, 0.917) | 0.741 (0.616, 0.891) |
| 12+ months | 0.884 (0.808, 0.967) | 0.864 (0.783, 0.953) | 0.884 (0.809, 0.967) | 0.882 (0.806, 0.965) | 0.882 (0.807, 0.964) |

In our primary analysis, we matched cases and controls in a 1:2 ratio and used a conditional logistic regression model to assess indirect protection. Here we ran separate sensitivity analyses to assess robustness of results to these analytical choices. We matched cases and controls in a 1:1 ratio, and we conducted the analysis without matching by time since most recent vaccine or infection, with additional statistical adjustment for time since most recent vaccine and/or infection, and without statistical adjustment for age and risk covariates in the conditional logistic regression model.

**Supplementary Table 15. Sensitivity analyses of regression model specification for indirect protection from vaccine and infection-acquired immunity.**

| | Odds ratio (OR) (95% CI) | | | |
|---|---|---|---|---|
| | **Conditional logistic regression (main)** | **Unconditional logistic regression** | **Conditional logistic regression without repeated residents** | **Unconditional logistic regression with person-level clustering** |
| **Vaccine-derived immunity** | | | | |
| Any | 0.778 (0.694, 0.872) | 0.800 (0.715, 0.896) | 0.799 (0.697, 0.916) | 0.800 (0.709, 0.904) |
| By dose | | | | |
| Partially vaccinated | 0.925 (0.891, 0.96) | 0.938 (0.904, 0.973) | 0.934 (0.893, 0.976) | 0.938 (0.902, 0.975) |
| Primary series only | 0.856 (0.824, 0.888) | 0.879 (0.848, 0.912) | 0.872 (0.833, 0.912) | 0.879 (0.845, 0.915) |
| One booster | 0.791 (0.763, 0.821) | 0.825 (0.795, 0.855) | 0.814 (0.778, 0.851) | 0.825 (0.793, 0.858) |
| Two or more boosters | 0.732 (0.705, 0.76) | 0.773 (0.745, 0.802) | 0.76 (0.726, 0.795) | 0.773 (0.743, 0.804) |
| By time | | | | |
| <3 months | 0.703 (0.619, 0.798) | 0.735 (0.650, 0.831) | 0.743 (0.638, 0.865) | 0.735 (0.644, 0.839) |
| 3-6 months | 0.871 (0.749, 1.01) | 0.878 (0.761, 1.01) | 0.834 (0.698, 0.998) | 0.878 (0.753, 1.02) |
| 6-12 months | 0.830 (0.727, 0.949) | 0.851 (0.748, 0.968) | 0.856 (0.73, 1.00) | 0.851 (0.740, 0.978) |
| 12+ months | 0.820 (0.671, 1.00) | 0.835 (0.689, 1.01) | 0.837 (0.661, 1.06) | 0.835 (0.683, 1.02) |
| **Infection-acquired immunity** | | | | |
| Any | 0.841 (0.773, 0.915) | 0.847 (0.781, 0.918) | 0.831 (0.752, 0.919) | 0.847 (0.776, 0.925) |
| By time | | | | |
| <3 months | 0.628 (0.451, 0.875) | 0.665 (0.481, 0.907) | 0.556 (0.368, 0.84) | 0.665 (0.479, 0.921) |
| 3-6 months | 0.616 (0.476, 0.796) | 0.667 (0.525, 0.843) | 0.535 (0.382, 0.748) | 0.667 (0.517, 0.861) |
| 6-12 months | 0.746 (0.619, 0.898) | 0.781 (0.658, 0.925) | 0.812 (0.645, 1.02) | 0.781 (0.652, 0.936) |
| 12+ months | 0.884 (0.808, 0.967) | 0.884 (0.811, 0.963) | 0.870 (0.782, 0.968) | 0.884 (0.805, 0.970) |

In our primary analysis, we used a conditional logistic regression model to assess indirect protection within groups of matched cases and controls. Here we assessed the sensitivity of results to potential bias from matching by fitting an unconditional logistic regression model with additional adjustment for factors that were exactly matched between cases and controls (vaccine and prior infection status of cases and controls and building and security level). To evaluate robustness of results to repeated observations from cases and controls, we fit a conditional logistic regression model without repeated measures and fit an unconditional logistic regression model with person-level cluster robust errors. Results across all models were similar.

### Supplementary Figures

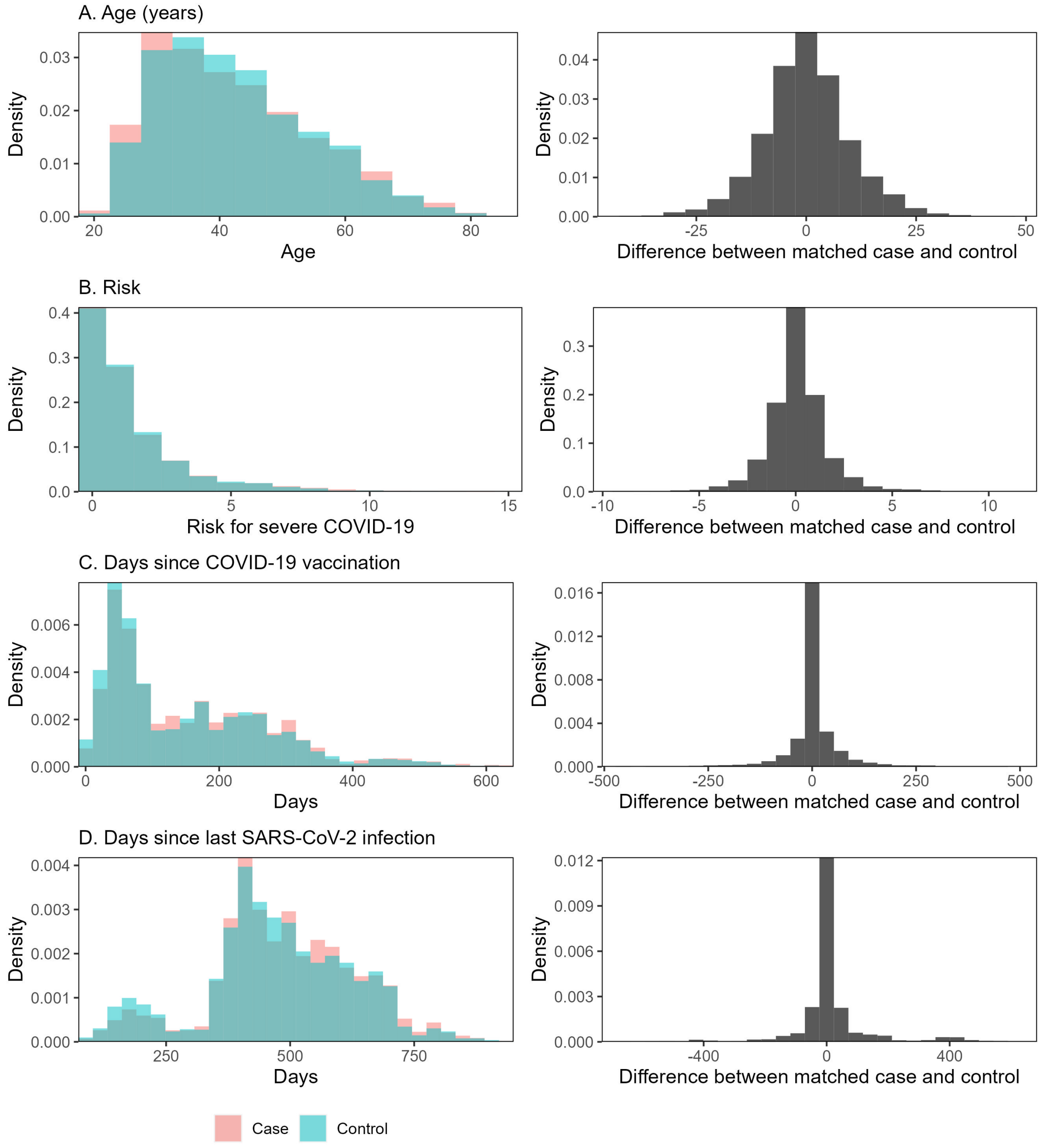


**Supplementary Figure 1. Match quality of matched cases and controls in study population.** We matched cases and controls exactly by building and security level, time, prior infection status (binary), and vaccine status (by dose). Cases and controls were then matched by distance on variables of age, severe COVID-19 risk score, time since last SARS-CoV-2 infection, and time since last COVID-19 vaccination. Here, we plotted the overall distributions of distance-matched variables of cases and controls on the left. On the right, we plotted differences between cases and controls within matched pairs. A positive difference favors cases, and a negative difference favors controls, though all difference distributions are symmetrical around 0.

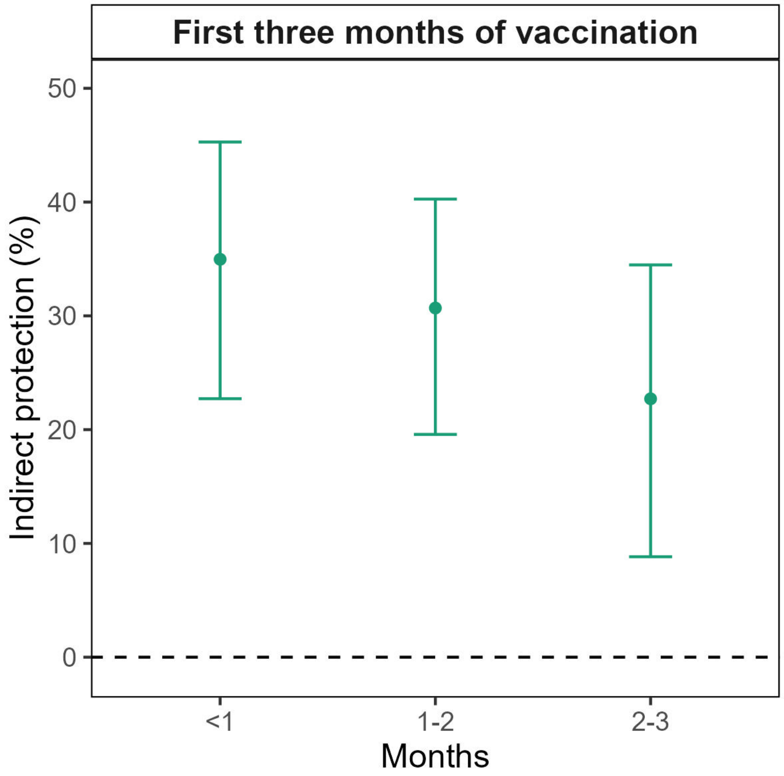


**Supplementary Figure 2. Vaccine-derived indirect protection within 3 months of vaccination.** We estimated the strength and durability of indirect protection that COVID-19 vaccination provided to their roommate in the first three months of COVID-19 vaccination. We defined onset of vaccine protection as 14 days after a dose. We plotted the mean (point estimate) and associated 95% confidence intervals (bars) for indirect protection.
